# A Phase IV, open-label, single-arm, multicentric clinical trial for evaluation of Human Papillomavirus 9vHPV vaccine immunogenicity in Men Who Have Sex with Men living with HIV: GeSIDA Study 10017

**DOI:** 10.1101/2025.05.08.25327237

**Authors:** Raquel Ron, Claudio Díaz-García, Elena Sendagorta, Alfonso Cabello-Úbeda, Elena Moreno, Clara Crespillo-Andújar, Rosa Feltes-Ochoa, Irene Carrillo-Acosta, Roser Navarro-Soler, Herminia Esteban, Miguel Górgolas, Santiago Moreno, José A. Pérez-Molina, Sergio Serrano-Villar

**Author notes:** **Corresponding authors:** Sergio Serrano-Villar, José A. Pérez-Molina. Department of Infectious Diseases. Ctra. de Colmenar Viejo, km 9.100. 28034 Madrid, Spain. RR and CDG contributed equally to this manuscript as co-first authors. JAPM and SSV contributed equally to this manuscript as co-senior authors.

## Abstract

**Background:** Men who have sex with men (MSM) with HIV are at increased risk for anal cancer, largely attributable to persistent HPV infection. However, data on the nonavalent HPV vaccine (9vHPV) in MSM with HIV older than 26 remain sparse.

**Methods:** This phase IV trial evaluated the immunogenicity, safety, and impact of age and CD4/CD8 ratio on 9vHPV responses in MSM with HIV up to 35 years. Inclusion criteria were age 16–35, undetectable viral load, and CD4+ counts >200 cells/mm^3^. Participants received 9vHPV at weeks 0, 8, and 24, and were followed to week 96. Serum samples for immunogenicity, anal HPV DNA testing, and adverse event reports were collected at baseline, week 28, and week 96. Additionally, we investigated potential predictors of vaccine immunogenicity, including age group and CD4/CD8 ratio.

**Results:** Among 157 enrolled participants, 138 completed the per-protocol analysis. Seroconversion exceeded 85% for all nine HPV vaccine genotypes at week 96. New infections with vaccine-included genotypes occurred in 39.9% of participants, predominantly involving HPV-16. Clearance of existing HPV infections covered by the 9vHPV was 73.8%, and highest (≥80%) for HPV-18. Neither older age (≥26 years) nor lower CD4/CD8 ratio significantly reduced immunogenicity. No severe adverse events related to vaccination were recorded.

**Conclusions:** The 9vHPV vaccine demonstrated robust immunogenicity and encouraging viral clearance rates in MSM with HIV up to 35 years. These findings support extending vaccination beyond 26 years in this high-risk group, emphasizing the additional coverage provided by the 9vHPV formulation.

**Clinical trial registration:** EudraCT number 2018-000215-24.

**Key points:** 9-valent HPV vaccination in MSM living with HIV up to 35 years shows good immunological response independent of age and CD4/CD8 value, with a good safety profile, suggesting benefit on incident infections and viral clearance.

## INTRODUCTION

Anal cancer has an incidence rate of 2.2 cases per 100,000 people in the European Union, accounting for nearly 9,901 new cases in 2022 (1). This disease disproportionately affects men who have sex with men (MSM), particularly those living with HIV, where the incidence is 85 per 100,000 person-years compared to 19 per 100,000 in MSM without HIV (2). Consequently, MSM with HIV represent one of the most vulnerable groups for anal cancer and could benefit greatly from both vaccination and screening.

Human Papillomavirus (HPV) infection plays a central role in the development of squamous cell carcinoma of the anus (SCCA) (3), with HPV genotypes 16 and 18 detected in the majority of cases (4). In MSM, the prevalence of anal high-risk HPV is around 41%, and this increases to 74% in MSM with HIV (5). Several clinical guidelines and National Health Systems recommend HPV vaccination for MSM with HIV up to 45 years of age, extrapolating efficacy data of HPV vaccine in women to prevent cervical cancer (6–8). However, the natural history of anal cancer differs substantially from that of cervical cancer. Moreover, data on the immunogenicity of the 9vHPV HPV vaccine (9vHPV) in MSM with HIV over 26 years of age remains scarce, which is crucial for informing clinical decision-making and public health strategies tailored to older adults with varying degrees of immune dysfunction.

Although previous randomized clinical trials (RCTs) support the safety and effectiveness of HPV vaccines in people living with HIV (PLHIV) (9), recent studies have questioned vaccine efficacy in older individuals, particularly those with advanced immunosuppression (10,11). Low CD4/CD8 T-cell ratios have been linked to suboptimal vaccine responses, suggesting that immune dysfunction may influence protective immunity (12,13). Age over 26 has also been associated with a diminished response to HPV vaccination among PLHIV (14). This may be due to immunosenescence, a decline in immune function with age, and the compounding effects of HIV-related immune dysfunction (15).

Thus, clarifying 9vHPV vaccine immunogenicity and its impact on new HPV infections and viral clearance in MSM with HIV is of paramount importance to guide anal cancer prevention efforts in this high-risk population. This gap in the literature is increasingly relevant given the broader recommendation of 9vHPV in MSM with HIV up to 45 years, due to its expanded coverage of oncogenic genotypes. Our study aims to assess the immunogenicity of 9vHPV in MSM with HIV and to explore how age and CD4/CD8 ratio might shape vaccine responses in this susceptible population.

## METHODS

### Study design and participants

We conducted a phase IV, multicenter, open-label, single-arm clinical trial to evaluate the immunogenicity and safety of the 9vHPV vaccine in MSM living with HIV, aged up to 35 years. Participants were recruited from HPV clinics specialized in high resolution anoscopy (HRA) and anal high-grade squamous intraepithelial lesions (HSIL) screening at three tertiary hospitals in Madrid, Spain: Hospital Universitario Ramón y Cajal, Hospital Universitario La Paz, and Fundación Jiménez Díaz.

Inclusion criteria required participants to be MSM living with HIV, aged 16 to 35 years, with undetectable viral load, CD4+ counts >200 cells/mm^3^ within the past six months, and a history of insertive or receptive anal intercourse. Exclusion criteria were history of anal cancer, prior HPV vaccination, or being over 35 years of age.

The study adhered to the International Principles of Good Clinical Practice and received approval from the Ethics Committee for Clinical Investigation at Ramón y Cajal Hospital. Informed consent was obtained from all participants before conducting any study procedures. This study is registered with EudraCT Number: 2018-000215-24.

### Study Populations and Study Outcomes

The primary outcome of the study was the immunogenicity of the 9vHPV vaccine, assessed by changes in seroprevalence and antibodies geometric mean concentration (GMC) ratio from baseline to weeks 28 and 96. The secondary outcomes included HPV prevalence, incidence, and clearance at each time point.

For the analysis, three distinct study populations were defined:

1. Per-protocol population: This group included participants who received all three doses of the 9vHPV vaccine as per the study protocol, had at least one serological assessment after the third dose (at week 28 or later), and without any protocol violations. Participants in this group were required to be seronegative at baseline for at least one vaccine-included genotype (or genotypes 6 and 11 simultaneously, if applicable).
2. Intention-to-treat population: This group comprised participants who received at least one dose of the 9vHPV vaccine and had at least one serological assessment at week 28 or later. Unlike the per-protocol population, participants in this group could be seronegative or seropositive for vaccine-included genotypes at baseline.
3. Safety population: This group included all participants who received at least one dose of the 9vHPV vaccine, regardless of serological assessments or protocol adherence, and were monitored for adverse events, toxicity, and laboratory safety parameters.

### Study Procedures and Laboratory Measurements

Participants received a 0.5 mL intramuscular injection of the 9vHPV vaccine on day 0, week 8, and week 24. At each visit, they received a vaccination report card to record any adverse events. Serum samples were collected at baseline, week 28, and week 96 for further analysis.

As part of routine clinical care provided at HPV clinics, participants underwent an anorectal digital examination at each visit. Additional procedures included HPV DNA testing, anorectal PCR testing for *Chlamydia trachomatis* and *Neisseria gonorrhoeae* infections, and a liquid-based anal cytology sample collected with a cytobrush. If cytological abnormalities were detected, an HRA was performed by a trained physician. Participants diagnosed with HSIL received treatment and follow-up following local standards of care.

Anal samples were collected using the Anex Brush (Rovers Medical Devices, Netherlands) and rinsed in 20 ml of PreservCyt fixative (Hologic, Inc., Marlborough, MA, USA). Cytology was evaluated by expert cytopathologists at each center and classified per the Bethesda System as unsatisfactory, negative, ASCUS, LSIL, ASC-HSIL, or HSIL (16). HPV testing was performed with the Allplex™ HPV28. This multiplex real-time PCR assay enables simultaneous amplification and detection of target nucleic acids of 19 high-risk HPV types (16, 18, 26, 31, 33, 35, 39, 45, 51, 52, 53, 56, 58, 59, 66, 68, 69, 73, 82) and 9 low-risk HPV types (6, 11, 40, 42, 43, 44, 54, 61, 70).

Plasma samples were analyzed for antibodies against HPV types 6, 11, 16, 18, 31, 33, 45, 52, and 58 using the Luminex competitive HPV immunoassay (HPV-9 cLIA) at baseline, week 28, and week 96.

### Statistical analysis

A sample size of 150 participants was calculated to address the study objectives, primarily to evaluate the impact of age and CD4/CD8 ratio on immunogenicity. This estimation was based on prior data on HBV and yellow fever vaccine immunogenicity in people living with HIV (PLHIV) (12,17,18). To account for a potential 10% loss to follow-up over the study period, the final target sample size was adjusted to include 166 participants.

A per-protocol analysis was performed to evaluate immunogenicity by age group. The seroconversion rate was defined as the proportion of participants who were seronegative for each HPV genotype at baseline and developed antibody concentrations above the manufacturer-defined cut-off values at weeks 28 and 96 (19). Geometric mean concentrations (GMC) were calculated as the nth root of the product of antibody concentrations, where n represents the sample size. Seroconversion rates 95% confidence intervals (CIs) were calculated using the Clopper-Pearson (exact binomial) method. GMCs 95% CIs were computed based on the normality assumption.

Prevalence, incidence, clearance, persistence, and a combined variable for anal HPV infection were calculated. Prevalence was established as the proportion of participants who were positive at week 0. Incidence was calculated as the proportion of participants with a baseline negative test who tested positive for vaccine genotypes after vaccination. Clearance was the proportion of patients with a positive baseline test who tested negative for any genotype after vaccination. Persistence was established as the proportion of participants with a baseline positive test who remained positive after vaccination. Finally, the combined variable for positivity was computed as the proportion of patients with persistent infection plus those with positive HPV DNA in the final visit who were free of infection at the initial time.

To assess differences in immunogenicity associated with age and CD4/CD8 ratio, we assumed a binomial distribution for the seroconversion status and a logarithmic normal distribution for the HPV antibody concentrations. Logistic regression was used to analyze the relationship between age and CD4/CD8 ratio and the likelihood of seroconversion. This approach allowed us to quantify how these predictors influenced the probability of participants developing antibodies for each HPV genotype. Similarly, linear regression was applied to explore the impact of these variables on antibody concentration levels. In these models, seroconversion status and antibody concentrations were dependent variables, while age and CD4/CD8 ratio were treated as independent variables. Analyses were performed with RStudio and R version 4.3 (R Core Team).

## RESULTS

### Participants Characteristics

Between October 2018 and August 2019, 158 participants were enrolled. Of these, 157 received at least one dose of the 9vHPV vaccine, 139 completed week 28, and 123 participants (78%) completed the final study visit. The discontinuations were attributed to loss of follow-up (76%), early withdrawal (12%), protocol violations (9%) and screening failure (3%). Antibody determination samples from 10 participants were unavailable, and these individuals were excluded from the analysis. Also, four participants had the second antibody sample collected less than one month after receiving the third vaccine dose and, therefore, were not included in the per-protocol population.

The per-protocol population comprised 138 patients, while the safety population included 157 enrolled participants. The study flow chart is detailed in **Figure 1**. Baseline characteristics are summarized in **Table 1**. Most participants were white, with a mean age of 31 years and a median ART duration of 4 years. The median CD4 T-cell counts and CD4/CD8 ratio were 726 cells/uL and 0.85, respectively.

**Figure 1.**
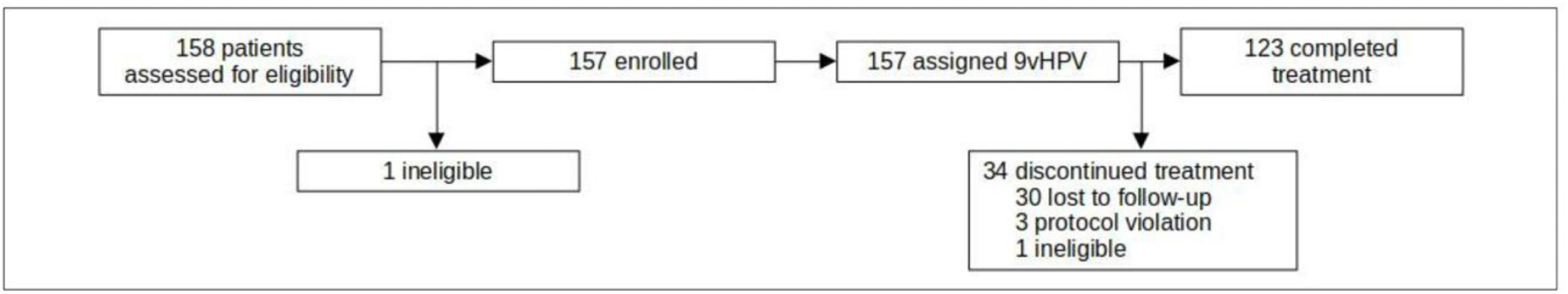
Participants flowchart.

**Table 1.** Baseline characteristics.

|  | Per-protocol population<br>(n = 138) | Safety population<br>(n = 157) |
| --- | --- | --- |
| Age (mean, (SD)) | 30.99 (3.54) | 30.88 (3.52) |
| Nadir CD4 | 409.25 (290.00, 599.85) | 412.00 (264.00, 600.00) |
| CD4 | 725.62 (553.50, 925.00) | 729.24 (562.50, 936.50) |
| CD4/CD8 | 0.85 (0.64, 1.07) | 0.84 (0.64, 1.07) |
| Years since HIV diagnosis | 4.00 (2.00, 8.00) | 4.00 (2.00, 8.00) |
| Number of sexual partners per month | 1.00 (1.00, 2.00) | 1.00 (1.00, 2.00) |
| **Ethnicity** |  |  |
| White | 125 (90.6%) | 141 (89.8%) |
| Latin | 10 (7.2%) | 13 (8.3%) |
| Black | 3 (2.2%) | 3 (1.9%) |
| **Smoking** |  |  |
| Yes | 56 (40.6%) | 64 (40.8%) |
| N/A | 1 (0.7%) | 1 (0.6%) |
| **Drug use in sexual context** |  |  |
| Yes | 22 (16.3%) | 26 (17.0%) |
| **HIV transmission** |  |  |
| MSM | 136 (98.6%) | 154 (98.7%) |
| Unknown | 2 (1.4%) | 2 (1.3%) |
| ***Chlamydia trachomatis*** |  |  |
| Positive | 7 (5.1%) | 11 (7.1%) |
| N/A | 2 (1.5%) | 2 (1.3%) |
| ***Neisseria gonorrhoeae*** |  |  |
| Positive | 9 (6.6%) | 11 (7.1%) |
| N/A | 3 (2.2%) | 3 (1.9%) |

Continuous variables (except age) are shown as median and IQR, while categorical variables are shown as frequency and percentages.

### Seroconversion

**Table 2** details the baseline prevalence of anal HPV infection with vaccine-covered genotypes. Among these, HPV16 exhibited the highest prevalence (22%), while non-16/18 oncogenic genotypes ranged from 11.6% to 16.7%. In contrast, HPV18 had the lowest prevalence, observed in 8.0% of participants.

**Table 2.** Prevalence of anal HPV infection and HPV antibodies measured by PCR and CLIA.

| HPV genotype | Participants with positive PCR (n = 138) | Prevalence (%) | Patients with seropositive determination (n = 138) | Prevalence (%) |
| --- | --- | --- | --- | --- |
| High risk | 76 | 55.9 | NA | NA |
| HPV6 and HPV 11 | NA | NA | 35 | 25.4 |
| HPV-16 | 30 | 21.7 | 58 | 42.0 |
| HPV-18 | 11 | 8.0 | 58 | 42.0 |
| HPV-31 | 22 | 15.9 | 40 | 29.0 |
| HPV-33 | 22 | 15.9 | 36 | 26.1 |
| HPV-45 | 16 | 11.6 | 27 | 19.6 |
| HPV-52 | 22 | 15.9 | 18 | 13.0 |
| HPV-58 | 23 | 16.7 | 42 | 30.4 |
| Other | 87 | 63.0 | NA | NA |

Total number and percentage of participants who tested positive for vaccine-covered genotypes at baseline before vaccination (PCR prevalence) and with an antibody determination higher than the seroconversion threshold determined by the manufacturer (antibody prevalence).

In participants with a baseline negative serologic determination for each of the nine vaccine genotypes, the seroconversion rate, determined by plasma-neutralizing antibody concentrations—was nearly 100% for all genotypes at week 28. This high seroconversion rate persisted above 85% for all genotypes at week 96 post-vaccination (**Table 3**). The average plasma antibody concentrations for each HPV genotype, expressed as GMC, are summarized in **Table 4**.

**Table 3.** Seroconversion rates at weeks 28 and 96.

| HPV genotype | Week 28 |  | Week 96 |  |
| --- | --- | --- | --- | --- |
|  | Participants<br>(n = 135) | Seroconversion<br>(%, IC<sub>95</sub>) | Participants<br>(n = 119) | Seroconversion<br>(%, IC<sub>95</sub>) |
| HPV-6 and HPV-11 | 101 | 99<br>(94.6 - 100) | 87 | 94.3<br>(87.1 - 99.1) |
| HPV-16 | 79 | 100<br>(95.4 - 100) | 71 | 95.8<br>(88.1 - 100) |
| HPV-18 | 77 | 98.7<br>(93.0 - 100) | 65 | 87.7<br>(77.2 - 95.7) |
| HPV-31 | 96 | 99.0<br>(94.3 - 100) | 81 | 85.2<br>(75.6 - 92.9) |
| HPV-33 | 100 | 99.0<br>(94.6 - 100) | 85 | 91.8<br>(83.8 - 97.6) |
| HPV-45 | 110 | 99.1<br>(95.0 - 100) | 97 | 87.6<br>(79.4 - 94.2) |
| HPV-52 | 119 | 99.2<br>(95.4 - 100) | 104 | 93.3<br>(86.6 - 98.1) |
| HPV-58 | 94 | 100<br>(96.2 - 100) | 79 | 93.7<br>(85.8 - 99.0) |

Seroconversion rate for vaccine HPV genotypes in the per-protocol population, measured as the proportion of patients with an antibody concentration below the minimum value threshold indicated by the manufacturer at baseline and above this threshold after vaccination.

**Table 4.** Geometric mean concentration (GMC) and GMC ratios at weeks 28 and 96.

| HPV genotype | Week 28 |  |  | Week 96 |  |  |
| --- | --- | --- | --- | --- | --- | --- |
|  | Participants (n = 135) | GMC (mMU/mL, IC<sub>95</sub>) | GMC Ratio | Participants (n = 119) | GMC (mMU/mL, IC<sub>95</sub>) | GMC Ratio |
| HPV-6 | 101 | 840.7<br>(689.0 - 1026.0) | 19.9<br>(15.7 - 25.4) | 87 | 249.1<br>(197.4 - 314.5) | 5.9<br>(4.5 - 7.7) |
| HPV-11 | 101 | 511.6<br>(431.9 - 605.9) | 33.1<br>(27.4 - 40.0) | 87 | 142.7<br>(114.5 - 178.0) | 9.2<br>(7.3 - 11.7) |
| HPV-16 | 79 | 1903.9<br>(1558.4 - 2326.0) | 117.0<br>(94.5 - 145.0) | 71 | 308.6<br>(242.7 - 392.5) | 19.0<br>(14.7 - 24.4) |
| HPV-18 | 77 | 588.9<br>(476.3 - 728.1) | 14.9<br>(12.0 - 18.7) | 65 | 123.0<br>(105.0 - 144.1) | 3.1<br>(2.6 - 3.7) |
| HPV-31 | 96 | 307.5<br>(255.7 - 369.8) | 23.0<br>(18.6 - 28.3) | 81 | 66.0<br>(55.5 - 78.3) | 4.9<br>(4.0 - 6.0) |
| HPV-33 | 100 | 252.2<br>(216.5 - 293.8) | 21.1<br>(17.8 - 25.0) | 85 | 55.3<br>(46.8 - 65.3) | 4.6<br>(3.9 - 5.6) |
| HPV-45 | 110 | 142.7<br>(118.3 - 172.0) | 18.8<br>(15.4 - 22.9) | 97 | 31.1<br>(26.9 - 35.9) | 4.1<br>(3.5 - 4.8) |
| HPV-52 | 119 | 256.9<br>(219.7 - 300.5) | 26.7<br>(22.5 - 31.7) | 104 | 62.1<br>(52.5 - 73.5) | 6.5<br>(5.4 - 7.8) |
| HPV-58 | 94 | 242.0<br>(205.5 - 284.9) | 33.4<br>(28.0 - 40.0) | 79 | 48.7<br>(40.7 - 58.1) | 6.7<br>(5.6 - 8.2) |

For each genotype, the geometric mean concentrations are calculated on the patients with antibody levels below the seroconversion threshold at baseline from the initial 138 participants. GMC ratios are related to GMC at week 0.

### HPV infection incidence, clearance, and persistence

The incidence of HPV infection was defined as a positive HPV DNA test at week 28 or week 96 after vaccination among participants who tested negative at baseline (**Table 5**). Among the genotypes, HPV-16 had the highest incidence (13.9%) at week 96, followed by HPV-58 (11.3%) and HPV-33 (9.5%). Conversely, HPV-18 exhibited the lowest incidence at 1.6%. The incidence of infection with any high-risk HPV genotype covered by the vaccine was 44.1% at week 28 and decreased to 39.9% at week 96. Due to variability in laboratory detection methods, the prevalence and incidence of genotypes 6 and 11 could not be determined.

**Table 5.** Incidence of HPV infection at weeks 28 and 96.

| HPV genotype | Week 28 |  |  |  | Week 96 |  |  |  |
| --- | --- | --- | --- | --- | --- | --- | --- | --- |
| | Participants with positive PCR | Participants tested | Incidence (%) | Incidence ($\times 1000$ p-m) | Participants with positive PCR | Participants tested | Incidence (%) | Incidence ($\times 1000$ p-m) |
| HR-HPV* | 36 | 57 | 63.2 | 90.3 | 31 | 49 | 63.3 | 26.4 |
| HPV-16 | 13 | 106 | 12.3 | 17.6 | 15 | 108 | 13.9 | 5.8 |
| HPV-18 | 7 | 125 | 5.6 | 8 | 2 | 127 | 1.6 | 0.7 |
| HPV-31 | 11 | 114 | 9.6 | 13.7 | 9 | 116 | 7.8 | 3.3 |
| HPV-33 | 12 | 114 | 10.5 | 15 | 11 | 116 | 9.5 | 4 |
| HPV-45 | 5 | 120 | 4.2 | 6 | 10 | 122 | 8.2 | 3.4 |
| HPV-52 | 11 | 114 | 9.6 | 13.7 | 8 | 116 | 6.9 | 2.9 |
| HPV-58 | 11 | 113 | 9.7 | 13.9 | 13 | 115 | 11.3 | 4.7 |
| Other | 16 | 51 | 31.4 | 44.9 | 20 | 51 | 39.2 | 16.3 |

Total number and percentage of participants who tested positive for vaccine genotypes at week 28 or 96 after vaccination, with a baseline negative test. \*HR-HPV includes infection with any of the 19 genotypes detected by PCR.

Clearance was defined as the proportion of participants with a baseline positive HPV DNA test who tested negative for HPV at week 28 and 96 after vaccination, or those who tested negative at week 96 (**Table 6**). By week 96, the overall clearance rate for any high-risk HPV genotype included in the vaccine was 63.1% at week 28, increasing to 73.8% at week 96, with the highest clearance observed for HPV-18 at 81.8%.

**Table 6:** Clearance of HPV infection at weeks 28 and 96.

| HPV genotype | Week 28 |  |  |  | Week 96 |  |  |  | Week 28 and 96 |  |  |  |
| --- | --- | --- | --- | --- | --- | --- | --- | --- | --- | --- | --- | --- |
|  | Negative PCR after vaccination | Baseline positive PCR | Clearance (%) | Clearance (× 1000 p-m) | Negative PCR after vaccination | Baseline positive PCR | Clearance (%) | Clearance (× 1000 p-m) | Negative PCR after vaccination | Baseline positive PCR | Clearance (%) | Clearance (× 1000 p-m) |
| HR-HPV* | 13 | 76 | 18.1 | 25.9 | 12 | 66 | 18.2 | 7.6 | 6 | 66 | 8.2 | 3.4 |
| HPV-16 | 9 | 30 | 30.0 | 42.9 | 16 | 30 | 53.3 | 22.2 | 5 | 30 | 16.7 | 7.0 |
| HPV-18 | 7 | 11 | 63.6 | 90.9 | 9 | 11 | 81.8 | 34.1 | 7 | 11 | 63.6 | 26.5 |
| HPV-31 | 11 | 22 | 50.0 | 71.4 | 12 | 22 | 54.5 | 22.7 | 7 | 22 | 31.8 | 13.2 |
| HPV-33 | 13 | 22 | 59.1 | 84.4 | 13 | 22 | 59.1 | 24.6 | 7 | 22 | 31.8 | 13.2 |
| HPV-45 | 12 | 16 | 75.0 | 107.1 | 11 | 16 | 68.8 | 28.7 | 9 | 16 | 56.2 | 23.4 |
| HPV-52 | 16 | 22 | 72.7 | 103.9 | 15 | 22 | 68.2 | 28.4 | 10 | 22 | 45.5 | 19.0 |
| HPV-58 | 14 | 23 | 60.9 | 87.0 | 15 | 23 | 65.2 | 27.2 | 10 | 23 | 43.5 | 18.1 |
| Other | 27 | 87 | 31.8 | 45.4 | 37 | 87 | 42.5 | 17.7 | 17 | 87 | 19.5 | 8.1 |

Total number and percentage of patients who were negative for any genotype at week 28, 96 or both after vaccination, with a positive baseline test. \*HR-HPV includes clearance of any of the 19 genotypes detected by PCR.

HPV persistence was defined as detecting the same HPV genotype at baseline and follow-up visits without evidence of clearance during the study period (**Table 7**). By week 96, the highest persistence was observed for HPV-16 at 46.7% and HPV-31 at 45.5%. The overall persistence rate for any high-risk HPV genotype included in the vaccine was 60% at week 28, decreasing to 44.6% at week 96.

**Table 7:** Persistence of HPV infection at weeks 28 and 96.

| HPV genotype | Week 28 |  |  | Week 96 |  |  |
| --- | --- | --- | --- | --- | --- | --- |
|  | Positive PCR after vaccination | Baseline positive PCR | Persistence (%) | Positive PCR after vaccination | Baseline positive PCR | Persistence (%) |
| HR-HPV* | 59 | 72 | 81.9 | 54 | 66 | 81.8 |
| HPV-16 | 21 | 30 | **70.0** | 14 | 30 | **46.7** |
| HPV-18 | 4 | 11 | 36.4 | 2 | 11 | **18.2** |
| HPV-31 | 11 | 22 | 50.0 | 10 | 22 | 45.5 |
| HPV-33 | 9 | 22 | 40.9 | 9 | 22 | 40.9 |
| HPV-45 | 4 | 16 | **25.0** | 5 | 16 | 31.2 |
| HPV-52 | 6 | 22 | 27.3 | 7 | 22 | 31.8 |
| HPV-58 | 9 | 23 | 39.1 | 8 | 23 | 34.8 |
| Other | 58 | 85 | 68.2 | 50 | 87 | 57.5 |

Total number and percentage of patients who were positive for any genotype at week 28 or 96 after vaccination, with a positive baseline test. \*HR-HPV includes persistence of any of the 19 genotypes detected by PCR.

We also calculated a combined variable, defined as the proportion of patients with persistent infection or detection of HPV DNA during the final visit of those free of infection at the initial time (**Supplementary Table 1**). HPV-16 and HPV-31 had the highest percentage for this combined variable, with 25% and 16.2% at week 28, respectively. At week 96, this percentage decreased to 21% and 13.8%, respectively. This combined variable decreased after week 96 for all vaccine genotypes except HPV-45 and HPV-58. The overall positivity rate of any vaccine-covered genotype, irrespective of initial detection, was 43.5% at week 28, which decreased to 39.9% at week 96.

### Cytologic impact

At baseline, 46.6% of participants had a normal cytology, maintaining a negative result at week 96, while 11.4% showed repeated abnormal cytology results during follow-up. At week 96, among patients with baseline normal cytology, 13.6% developed any cytologic abnormality. Of those with a baseline abnormal cytology, 17.1% transitioned to a negative result. The flow of changes between cytological categories is represented in the Sankey diagram (**Figure 2**).

**Figure 2.**
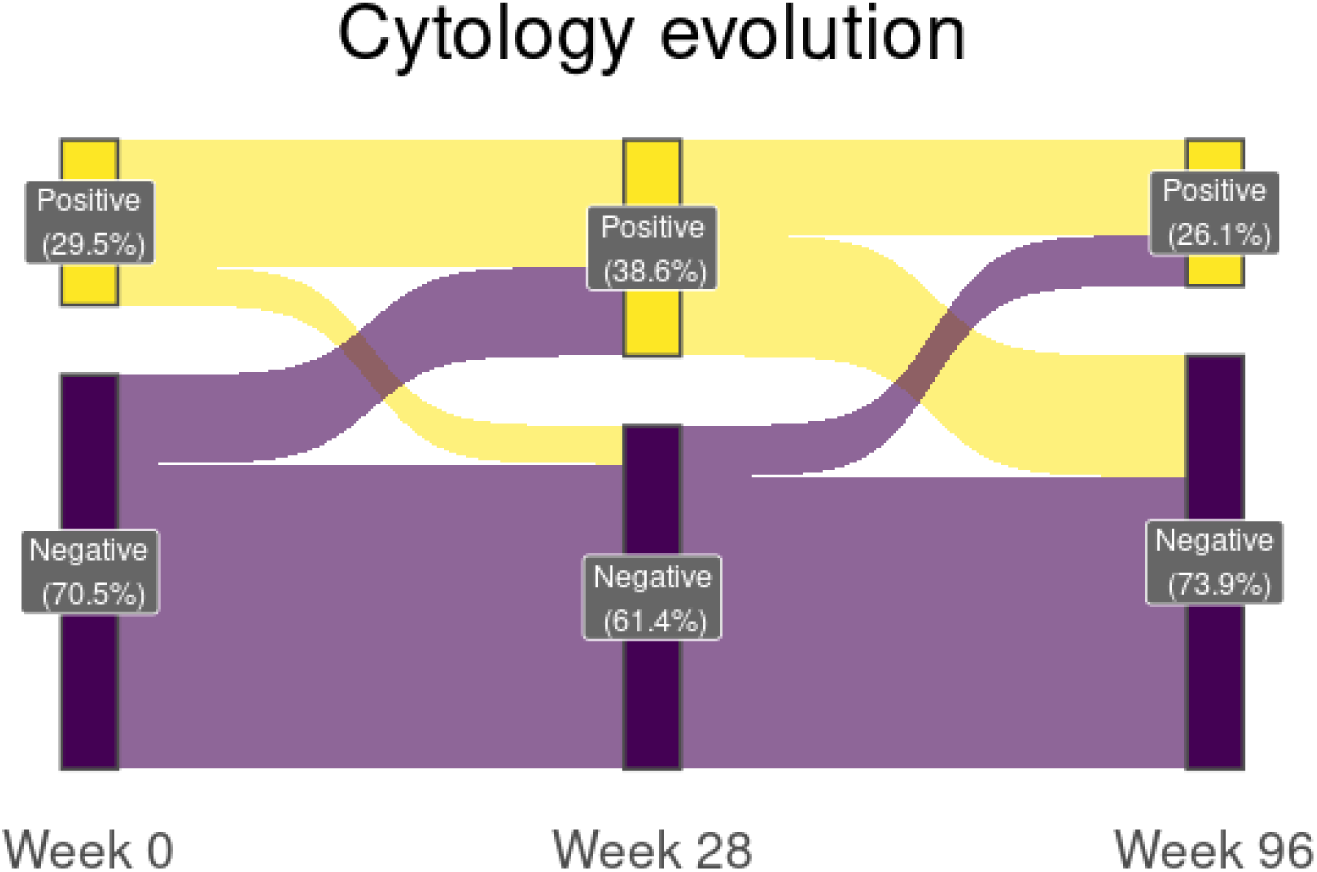
Sankey diagram of cytology results during follow-up. Positive results include those with ASCUS (Atypical Squamous Cells of Undetermined Significance), LSIL (Low-grade Squamous Intraepithelial Lesion), HSIL (High-grade Squamous Intraepithelial Lesion), or ASC-HSIL (combination of ASCUS and HSIL).

### Predictors of immunogenicity

We assessed the impact of age and immune status on vaccine efficacy. Age was analyzed as a continuous variable and showed no significant effect on the seroconversion rate or antibody concentration for any HPV genotype, except HPV-31 at week 28 for the antibody concentration [coefficient: −0.039, IC95: (−0.078, −0.001)] (**Supplementary Tables 2 and 3**). To further explore this potential effect, we conducted a secondary analysis stratifying participants by age quartiles, where no differences could be observed (**Figure 3**). Also, we show the antibody concentration distribution in patients younger than 26 years old vs. older, as this was the age limit recommendation by guidelines at the time of inclusion (**Supplementary Figure 1**). The CD4/CD8 ratio was also evaluated as a continuous variable and similarly showed no significant effect on seroconversion rates or antibody concentrations for any HPV genotype (**Supplementary Tables 4 and 5**). The antibody concentrations across CD4/CD8 ratio quartiles are shown in **Figure 4**.

**Figure 3.**
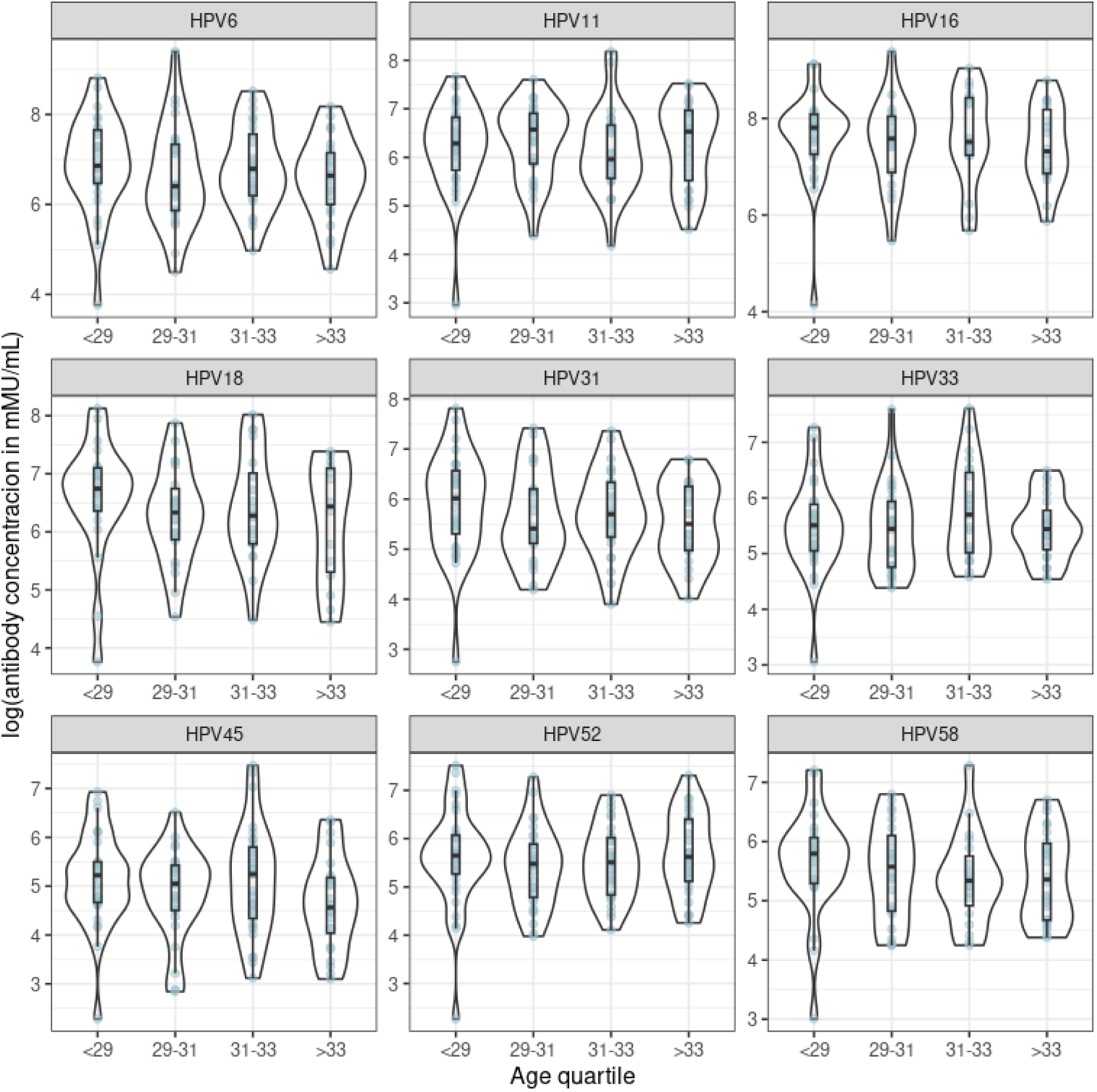
Antibody concentration at week 28 (4 weeks after vaccination) for each HPV genotype. Each group on the horizontal axis corresponds to a quartile of the age distribution.

**Figure 4.**
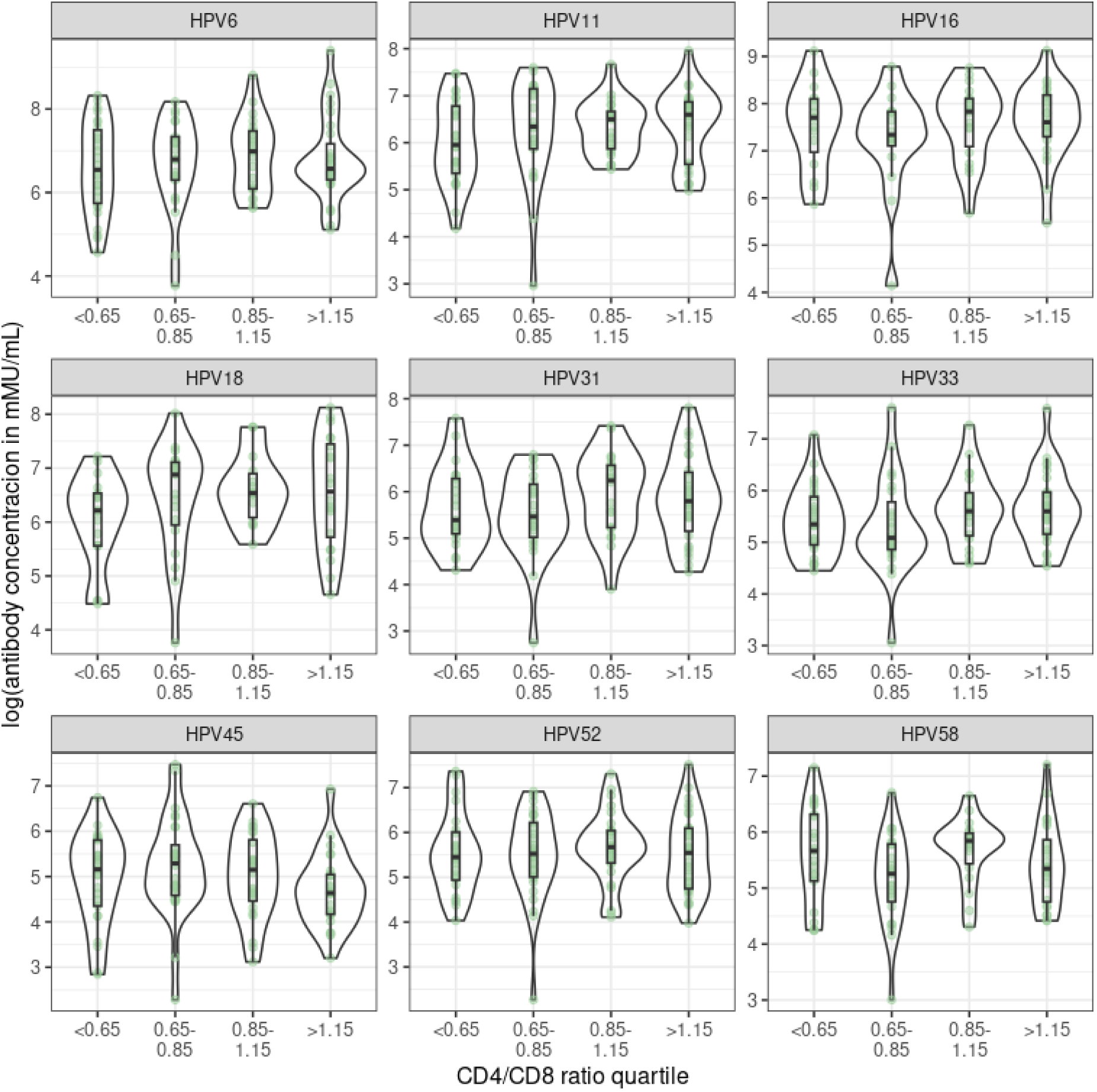
Antibody concentration at week 28 (4 weeks after vaccination) for each HPV genotype. Each group corresponds to a quartile of the CD4/CD8 ratio distribution.

### Safety

Adverse events reported during the study are summarized in **Supplementary Table 6**. While a high proportion of participants experienced at least one adverse event (68.8%), only a small percentage (1.9%) were deemed related to vaccination. The most frequently reported adverse events were local discomfort at the injection site and asthenia, which were mild in intensity and did not require study withdrawal or protocol modifications (**Supplementary Table 7**). No serious adverse events related to vaccination or deaths were recorded during the study.

## DISCUSSION

In this phase IV clinical trial conducted in MSM with HIV up to 35 years of age, we observed robust immunogenicity of the 9vHPV vaccine, with seroconversion remaining above 85% for all genotypes two years post-vaccination. These findings align with prior studies demonstrating strong vaccine-induced immune responses in older MSM with HIV who received quadrivalent HPV vaccination (20).

Notably, HPV-16 was the most prevalent genotype at baseline and displayed the highest incidence of new infection at week 96. We also found a higher prevalence of non-16/18 HPV infection, underscoring the relevance of these genotypes, being frequently detected in PLHIV (21). HPV-16 showed the highest persistence after vaccination (46.7%), mirroring its well-documented persistence in cohorts of MSM living with HIV. (21–23).

Despite high seroconversion rates, new high-risk HPV infections occurred in 63.3% of participants over two years, including infection with any of the 19 high-risk genotypes detected by PCR. This proportion likely reflects a selection bias stemming from recruitment in specialized HPV clinics, where participants often present with abnormal anal cytology or known high-risk HPV infection.

At week 96 after vaccination, we observed an incident rate of high-risk HPV of 26.4 x1000 persons-month (PM), and an incident rate for HPH-16 of 5.8 x1000 PM. Previous data in unvaccinated MSM living with HIV reported an incident rate of 36.1 and 10.2 x 1000 PM for high-risk HPV and HPV-16, respectively (24). In another cohort from the Netherlands, the incidence rate for HPV-16 in MSM living with HIV was 9.1 x 1000 PM (25). Our results support the benefit obtained through vaccination over incident infection, and the value of 9vHPV vaccination in this population.

The clearance rate at week 96 after vaccination was 7.6 x 1000 PM for any high-risk HPV, and 22.2 and 34.1 x 1000 PM, respectively, for HPV-16 and 18. In this scenario, the impact of vaccination is more controversial, as previous data in unvaccinated cohorts have reported clearance rates for any high-risk HPV and HPV 16 of 15.6 and 30.1 x 1000 pm, respectively (24). Encouragingly, we detected a 73.8% clearance rate for any high-risk HPV serotype covered by the vaccine, particularly against HPV-18, where clearance exceeded 80% by week 96. The reduced clearance of HPV-16 compared to HPV-18 is consistent with previous studies in the unvaccinated population (24,26,27), probably related to its higher oncogenic potential and persistent capacity, especially in PLHIV.

We also evaluated whether age or CD4/CD8 ratio influenced vaccine-induced seroconversion or antibody concentration. Previous data on MSM without HIV up to 45 years of age reveal vaccine effectiveness, especially in those vaccinated before 18 years or more than two years before specimen collection, but less so in those vaccinated over 26 years old (28). A meta-analysis likewise indicated limited vaccine effect in HIV-positive individuals older than 26 (14), although one phase III trial did demonstrate a modest 22% reduction in persistent infection after a median of 3.4 years (29).

In contrast to earlier reports suggesting reduced vaccine responses in those with an inverted ratio (13), and reduced efficacy on incidence and prevalence of HPV infection in older individuals (14), our data showed no significant differences in immunogenicity and seroconversion. Although this could be due to the relatively restricted age range (up to 35 years) and stable immune status (CD4 >200 cells/mm^3^, undetectable viral load) of our cohort, it nevertheless supports meaningful immunogenicity in MSM with HIV beyond the previous cutoff of 26 years.

Several limitations merit consideration. Our single-arm design precludes direct comparisons with unvaccinated controls, and the relatively small sample size may limit the detection of nuanced age or immune status-related differences. Also, the qualitative PCR test used for HPV detection is unable to distinguish between active or latent infection, and a negative result may reflect a low viral load instead of a complete viral clearance after vaccination. Larger, randomized studies are needed to validate these findings, evaluate vaccine impact on HSIL prevention at older age, and explore how behavioral factors, ART duration, and microbial signatures might shape long-term vaccine efficacy.

In conclusion, our data suggest that 9vHPV vaccination in MSM with HIV extends immunogenic benefits beyond 26 years of age, facilitating clearance of oncogenic HPV genotypes and contributing to the reduction of new infections. Given the substantial burden of anal cancer in this population, these findings reinforce current recommendations to vaccinate older age groups and emphasize the added coverage offered by the 9vHPV vaccine. Future research should incorporate detailed immunologic assessments and metagenomic approaches to identify host and microbial determinants of optimal vaccine response and disease prevention in older MSM with HIV.

## Data Availability

All data produced in the present study are available upon reasonable request to the authors.

## ACKNOWLEDGMENTS

“We thank all the patients and healthcare workers who participated in the study. This work was supported by Merck Sharp & Dohme [IISP 56519]; Instituto de Salud Carlos III (FI22/00111 to C.D.G.; projects ICI20/00058, PI21/00041, and PI24/00078 to SSV) and the European Union (NextGenerationEU); and was monitored by Fundación SEIMC-GESIDA [GESIDA 10017]. The funder had no role in the study design, data collection, data analysis, data interpretation, or manuscript writing.

## CRediT authors statement

R.R.: Methodology, Investigation, Writing - original draft

C.D.G.: Methodology, Validation, Formal Analysis, Investigation, Data curation, Writing - original draft, Visualization

E.S.: Investigation, Resources, Project administration A.C.: Investigation, Resources, Project administration E.M.: Methodology, Investigation

C.C.A.: Investigation R.F.O.: Investigation I.C.A.: Investigation R.N.: Investigation

H.E.: Investigation, Project administration M.G.: Investigation

S.M.: Investigation

J.A.P.M: Conceptualization, Methodology, Validation, Formal Analysis, Investigation, Resources, Writing - review & editing, Supervision, Project administration, Funding acquisition

S.S.V.: Conceptualization, Methodology, Validation, Formal Analysis, Investigation, Resources, Writing - review & editing, Supervision, Project administration, Funding acquisition

All authors have approved the final version of the manuscript, and we declare no conflicts of interest.

