## Supplementary materials for "A Phase IV, open-label, single-arm, multicentric clinical trial for evaluation of Human Papillomavirus 9vHPV vaccine immunogenicity in Men Who Have Sex with Men living with HIV: GeSIDA Study 10017"

**Key points**: 9-valent HPV vaccination in MSM living with HIV up to 35 years shows good immunological response independent of age and CD4/CD8 value, with a good safety profile, suggesting benefit on incident infections and viral clearance.

### SUPPLEMENTARY FIGURES

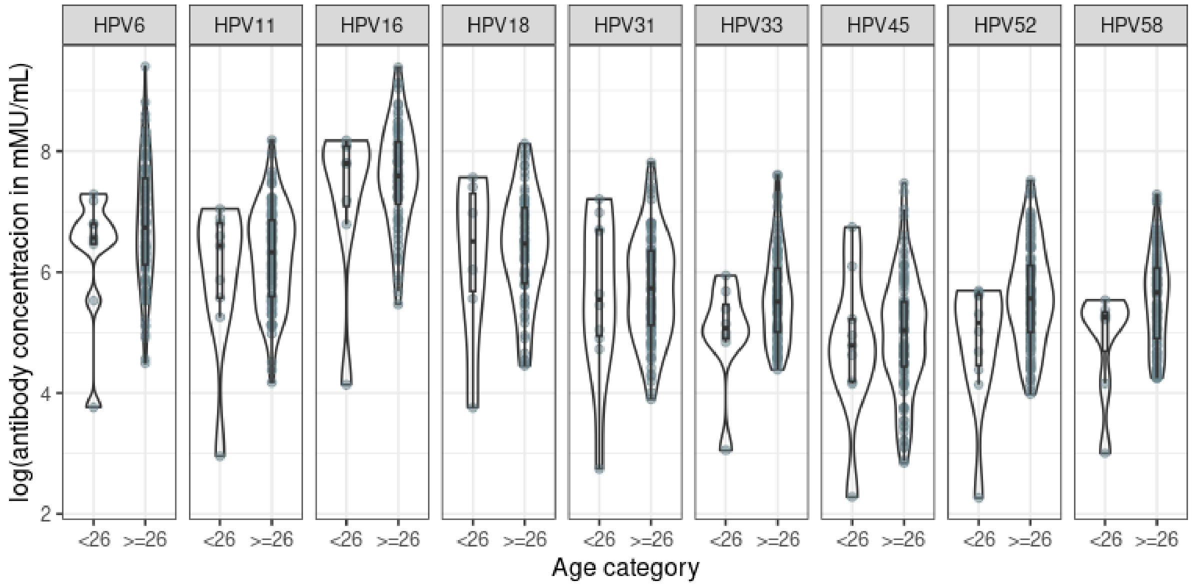


**Supplementary Figure 1**: Antibody concentration at week 28 (4 weeks after vaccination) for each HPV genotype. Each group on the horizontal axis corresponds to an age group, as depicted.

### SUPPLEMENTARY TABLES

**Supplementary Table 1.** Combined variable for positivity of HPV infection at weeks 28 and 96.

| HPV genotype | Week 28 | | | Week 96 | | |
|---|---|---|---|---|---|---|
| | Positive PCR after vaccination | Baseline positive PCR | Combined (%) | Positive PCR after vaccination | Baseline positive PCR | Combined (%) |
| **HR-HPV*** | 97 | 131 | 74.0 | 86 | 116 | 74.1 |
| **HPV-16** | 34 | 136 | 25.0 | 29 | 138 | 21.0 |
| **HPV-18** | 11 | 136 | 8.1 | 4 | 138 | 2.9 |
| **HPV-31** | 22 | 136 | 16.2 | 19 | 138 | 13.8 |
| **HPV-33** | 21 | 136 | 15.4 | 20 | 138 | 14.5 |
| **HPV-45** | 9 | 136 | 6.6 | 15 | 138 | 10.9 |
| **HPV-52** | 17 | 136 | 12.5 | 15 | 138 | 10.9 |
| **HPV-58** | 20 | 136 | 14.7 | 21 | 138 | 15.2 |
| **Other** | 74 | 136 | 54.4 | 70 | 138 | 50.7 |

Total number and percentage of patients with persistent infection or detection of HPV DNA in the final visit of those free of infection at baseline. *HR-HPV includes infection with any of the 19 genotypes detected by PCR.

**Supplementary Table 2**: Association between seroconversion and age at weeks 28 and 96.

| HPV serotype | Week 28 | | Week 96 | |
|---|---|---|---|---|
| | OR | IC95 | OR | IC95 |
| **HPV-6** | 1.140 | (1.172, 52.460) | 0.962 | (0.713, 1.221) |
| **HPV-11** | 1.140 | (1.172, 52.460) | 1.056 | (0.817, 1.324) |
| **HPV-16** | - | - | 1.003 | (0.705, 1.336) |
| **HPV-18** | 1.130 | (1.148, 47.911) | 0.938 | (0.735, 1.152) |
| **HPV-31** | 1.138 | (1.168, 52.665) | 0.836 | (0.660, 1.012) |
| **HPV-33** | 1.140 | (1.177, 49.739) | 0.987 | (0.772, 1.213) |
| **HPV-45** | 1.144 | (1.190, 49.472) | 0.805 | (0.616, 0.997) |
| **HPV-52** | 1.147 | (1.195, 52.894) | 1.028 | (0.816, 1.258) |
| **HPV-58** | - | - | 0.956 | (0.696, 1.239) |

In cases where patient seroconversion was 100%, it was not possible to compute these models (indicated by a dash in the coefficient table).

**Supplementary Table 3:** Association between antibody concentration and age at weeks 28 and 96.

| HPV serotype | Week 28 | | Week 96 | |
|---|---|---|---|---|
| | Coefficient | IC95 | Coefficient | IC95 |
| **HPV-6** | -0.003 | (-0.059, 0.052) | -0.013 | (-0.078, 0.052) |
| **HPV-11** | 0.017 | (-0.031, 0.064) | 0.020 | (-0.042, 0.081) |
| **HPV-16** | 0.003 | (-0.051, 0.057) | 0.008 | (-0.056, 0.073) |
| **HPV-18** | -0.003 | (-0.065, 0.058) | 0.009 | (-0.036, 0.054) |
| **HPV-31** | -0.023 | (-0.075, 0.028) | -0.038 | (-0.084, 0.009) |
| **HPV-33** | 0.016 | (-0.027, 0.060) | 0.029 | (-0.018, 0.077) |
| **HPV-45** | -0.026 | (-0.081, 0.029) | 0.001 | (-0.042, 0.044) |
| **HPV-52** | 0.024 | (-0.020, 0.069) | 0.026 | (-0.023, 0.074) |
| **HPV-58** | -0.007 | (-0.054, 0.041) | -0.010 | (-0.064, 0.043) |

**Supplementary Table 4**: Association between seroconversion and CD4/CD8 ratio at weeks 28 and 96.

| HPV serotype | Week 28 | | Week 96 | |
|---|---|---|---|---|
| | OR | IC95 | OR | IC95 |
| **HPV-6** | 1.564 | (0.033, 1369.872) | 1.393 | (0.199, 17.327) |
| **HPV-11** | 1.564 | (0.033, 1369.872 | 2.406 | (0.290, 39.306) |
| **HPV-16** | - | - | 27.489 | (0.658, 5292.474) |
| **HPV-18** | 1.730 | (0.023, 2225.180) | 9.866 | (0.900, 227.432) |
| **HPV-31** | 1.777 | (0.035, 2360.037) | 0.525 | (0.135, 2.222) |
| **HPV-33** | 1.524 | (0.033, 1569.069) | 0.679 | (0.144, 4.204) |
| **HPV-45** | 1.393 | (0.030, 1481.297) | 0.513 | (0.135, 2.192) |
| **HPV-52** | 1.805 | (0.030, 2242.766) | 0.700 | (0.136, 5.012) |
| **HPV-58** | - | - | 1.112 | (0.136, 14.143) |

In cases where patient seroconversion was 100%, it is not possible to compute these models (indicated by a dash in the coefficient table).

**Supplementary Table 5:** Association between antibody concentration and CD4/CD8 ratio at weeks 28 and 96.

| HPV serotype | Week 28 | | Week 96 | |
|---|---|---|---|---|
| | Coefficient | IC95 | Coefficient | IC95 |
| **HPV-6** | 0.254 | (-0.230, 0.738) | 0.091 | (-0.458, 0.640) |
| **HPV-11** | 0.203 | (-0.206, 0.612) | 0.132 | (-0.383, 0.648) |
| **HPV-16** | 0.069 | (-0.427, 0.564) | -0.116 | (-0.725, 0.492) |
| **HPV-18** | 0.438 | (-0.111, 0.986) | 0.084 | (-0.326, 0.494) |
| **HPV-31** | 0.171 | (-0.288, 0.629) | 0.066 | (-0.352, 0.484) |
| **HPV-33** | 0.196 | (-0.165, 0.557) | 0.016 | (-0.373, 0.405) |
| **HPV-45** | -0.200 | (-0.672, 0.272) | -0.284 | (-0.637, 0.069) |
| **HPV-52** | 0.038 | (-0.374, 0.451) | 0.016 | (-0.408, 0.439) |
| **HPV-58** | -0.030 | (-0.436, 0.375) | -0.157 | (-0.586, 0.271) |

**Supplementary Table 6**. Adverse events reported during the study.

| **Summary of patients with adverse events** | |
|---|---|
| Proportion of patients reporting an adverse event | 108 (68.8%) |
| Proportion of patients reporting an adverse event related to vaccination | 3 (1.9%) |
| Local discomfort after vaccination | 2 (<1%) |
| Asthenia | 1 (<1%) |
| Proportion of patients reporting a serious adverse event related to vaccination | 0 (0%) |
| Deaths | 0 (0%) |

**Supplementary Table 7.** Summary of adverse events reported during the study.

| **Summary of adverse events** | |
|---|---|
| Number of total adverse events | 303 |
| Severity of adverse effects | |
| Light | 281 (92,7%) |
| Moderate | 15 (5%) |
| Severe | 6 (2%) |
| Potentially life threatening | 1 (<1%) |
| Actions taken regarding the study vaccination | |
| None | 303 (100%) |
| Other actions taken | |
| None | 85 (28,1%) |
| Concomitant treatment | 214 (70,6%) |
| Procedures performed | 7 (2,3%) |
| Withdrawal of the patient from the study | 0 (0%) |
